# The genetic architecture of the β-globin chain in individuals with and without sickle cell disease in Nigeria: A case for beta thalassemia?

**DOI:** 10.1101/2024.12.20.24319473

**Authors:** Oluwatoyin Aduke Babalola, Foluke Atinuke Fasola, Biobele Jotham Brown, Jing Zhang, Yonglan Zheng, Abayomi Odetunde, Adeyinka Gladys Falusi, Olufunmilayo Olopade

**Affiliations:** Institute for Advanced Medical Research and Training, College of Medicine, University of Ibadan, Nigeria; Department of Hematology, College of Medicine, University of Ibadan, Nigeria; Department of Pediatrics, College of Medicine, University of Ibadan, Nigeria; Department of Medicine, University of Chicago, IL, USA; Department of Molecular Biology & Biotechnology, Chrisland University, Abeokuta, Nigeria

## Abstract

**Background:** Reports on beta thalassemia in the Nigerian population are conflicting, and the prevalence and role of beta thalassemia in Nigerian sickle cell disease (SCD) patients remain unclear. There is a need to set the records straight to ascertain the prevalence and effect of beta thalassemia in these patients.

**Methods:** 123 SCD patients and 117 age- and sex-matched controls were recruited. For the cases, the age range was 3 – 69 years, median(IQR) = 16(9 - 29). A separate cohort of 26 SCD patients were studied along. Full blood count was calculated, and the hemoglobin fractions were estimated by High Performance Liquid Chromatography. The 1.6kb beta-globin gene region was amplified from germline DNA and Sanger sequencing was performed. Single nucleotide polymorphisms (SNPs) were detected and annotated, and haplotypes were constructed.

**Results:** Contrary to recent reports, the prevalence of beta thalassemia in this population is <1%. Aside the sickle, hemoglobin C and beta thalassemia mutations, eight other variants were identified. Only three of these variants were found in the SCD patients and are in linkage disequilibrium. The sickle and hemoglobin C mutations arose on the major ancestral haplotype, while the beta thalassemia intermedia mutation (rs33915217C>A) was found on the minor ancestral haplotype, atypical of Africa. Two rare variants (rs537944366T>C and rs33915217C>A) are reported for the first time in the Yoruba population.

**Conclusion:** There is a need for a re-assessment of the diagnosis of beta thalassemia in this and other African populations for the proper management of SCD and other anemia-related cases.

## INTRODUCTION

Beta thalassemia is usually due to point mutations and occasionally deletions and insertions in the beta-globin gene locus^1^. The human beta-globin gene is part of a beta-globin cluster on the short arm of chromosome 11^2^. The beta globin gene consists of three exons and three introns, spanning about 1.6kb^2^. The gene is very diverse with as many as about 954 variants, about 300 or more of which result in beta thalassemia^3^. Beta thalassemia results from a reduction or lack of production of the beta-globin protein chain from the beta globin gene, which leads to a reduced production of adult hemoglobin (HbA)^4,5^. The reduced production in HbA causes anemia, but with varying levels of severity depending on the type of mutation and the type of inheritance, in the heterozygous or homozygous state^5^. There are three types of beta thalassemia - beta thalassemia minor, beta thalassemia intermedia and beta thalassemia major^6^.

Beta thalassemia occurs worldwide; regions that used to be free of the disease are now recording cases of beta thalassemia due to migration and inter-ethnic/inter-racial marriages^6,7^. However, beta thalassemia was endemically prevalent in the Mediterranean region, the Middle East and South East Asia with the lowest prevalence in North America and Northern Europe^7^. Cyprus, Sardinia and South East Asia have the highest carrier frequency of the disease ^8^. Global carrier frequency is about 1.5%^6^. Beta thalassemia was not common in the Western part of Africa, but previous reports^9,10^ have described a prevalence of beta thalassemia minor or beta thalassemia trait (β^+^-thal) of about 25% in the Nigerian general population. These reports were based on High Performance Liquid Chromatography (HPLC) measurements of HbA_2_ > 3·5% and HbF > 1%. However, Esan et al^11^ reported a frequency of <1%, using the starch-block electrophoresis method and a measurement of HbA_2_ > 3·8%.

Sickle cell disease (SCD) is also due to a point mutation on the same beta-globin gene and may be concurrently inherited with beta-thalassemia. SCD patients inheriting beta thalassemia concurrently may have a severe or milder phenotype, depending on the kind of beta thalassemia mutation^12,13^. A study based on the results of HPLC has reported a prevalence of 10·6% among Nigerian SCD patients^14^. A study that screened children five years and below in Northern Nigeria has reported a frequency of 0·4% of beta thalassemia based on molecular techniques, among children with SCD and none among children without SCD^15^. The diagnosis of beta thalassemia trait is mostly based on elevated HbA_2_, but other factors such as alpha thalassemia^16^, and the use of certain drugs like antiretrovirals, vitamin B12/folate deficiency and hyperthyroidism can also cause elevated levels of HbA_2_ in the blood^17^. Alpha thalassemia is very common in Nigeria at a prevalence above 35%^16,18^. Nigeria has the highest burden of SCD in the world, and the prevalence and effect of beta thalassemia in these patients are unclear. This study was carried out to ascertain the prevalence and genetic spectrum of beta-thalassemia in individuals with and without SCD and the possible interaction between beta thalassemia and SCD in Nigeria.

## MATERIALS AND METHODS

### Study design

This is a cross sectional study of 123 SCD patients in steady state, attending the University College Hospital’s Pediatric Outpatient Clinic and Hematology Clinic in Ibadan, Nigeria. 117 age- and sex-matched apparently healthy controls from the community were also recruited. Venous blood was collected from each participant and 3mL was added to EDTA bottles for complete blood count, high-performance liquid chromatography (HPLC), and DNA extraction. More details on the recruitment of study participants, demographic and clinical parameters; sampling, laboratory analyses and DNA extraction from whole blood have been previously described ^16^. A subset of twenty-six individuals (from another health facility, Ring-Road State Hospital, Ibadan) with peculiar hematological parameters was studied along with this cohort.

### Ethical approval

This study was approved by the University of Ibadan/University College Hospital, Ibadan Institutional Ethics Review Committee, College of Medicine, University of Ibadan, and the Institutional Review Board at the University of Chicago. Written consent form was obtained from each participant. The design and implementation of the research protocol were in accordance with the principles of The Declaration of Helsinki.

### PCR amplification of the beta-globin gene

The human beta-globin gene is located on chromosome 11p15.4, GRCh38:5225464 – 5227071. It is 1608 base pairs (bp) long and contains three exons and two introns^2^. This region was amplified in three separate PCR reactions using three different sets of primers (Supplementary Table 1). The regions amplified by each set of primers are shown in Supplementary Figure 1. Each 15µL PCR reaction contained 200µM of each dNTP, 1.5µL of 10x PCR buffer, 2.5mM MgCl_2,_ Taq polymerase, 1.5 µL of 20ng/µL of DNA template and 10µM of forward and reverse primers. PCR reactions were conducted in a thermocycler (PTC-200 Peltier Thermal Cycler, MJ Research) with an initial denaturation step at 98°C for 2 mins, followed by 35 cycles of 98°C denaturation for 45 secs, 63°C annealing temperature for 45 secs, and 72°C extension for 1 min 30 secs. A final 10-minute extension at 72°C was used to complete the reaction. Five microliters of each amplicon were analyzed by electrophoresis on a 1.5 % agarose gel in Tris-Acetic Acid-EDTA buffer at 120 volts for 45 mins. DNA samples from twenty-six (26) SCD patients having peculiar HPLC results (elevated HbF, >10% and/or elevated HbA_2_, >3%) in another study cohort from a different health facility were sequenced along with samples in this study. Only the first and third set of primers were used on this set of samples.

### Cleaning of beta-globin gene amplicons for sequencing

Amplicons were cleaned prior to sequencing. Each 10µL reaction contained 1µL SAP Buffer (10x), 0.5µL of 1U/µL SAP, 0.1µL of 20U/µL Exo 1, 3.4µL of PCR grade water and 5µL of amplicons. This was then incubated at 37°C for 45 mins, 95°C for 15 mins and 4°C forever. 4µL of the cleaned amplicons were submitted for Sanger sequencing.

### Analysis of amplicon sequences

The sequences of the 1.6kb beta-globin gene amplicons were read and SNPs were detected using the sequencher software version 4.2 (Gene Codes Corporation, Ann Arbor, MI USA). The reference sequence of the GRCh38·p14 assembly with accession number NG_059281.1 was used as a template for sequence comparisons. The HbVar database^3^ was used to determine the rs numbers of the SNPs found within the amplicons and dbSNP^19^, ClinVar,^20^ Ensembl Variation database^21^ were used to determine the significance of each of the SNPs and allele frequencies in Africa and across the globe.

### Haplotype analysis

Using Haploview version 4.2^22^, the ten SNPs detected in the healthy control group were analysed in one haplotype block since the full length of the amplicon is less than 2kb. Haploview was used to determine haplotype frequencies and SNPs in linkage disequilibrium within the beta-globin gene.

### Statistical analysis

Allele frequencies of SNPs detected in this study were compared with those found in the Ensembl 1000 Genomes Phase 3 project using student’s t-test.

## RESULTS

### PCR amplicons of the beta-globin gene

Three different sets of amplicons of size 774bp, 645bp and 574bp were obtained from the PCR reactions (Supplementary Figure 2).

### SNPs in the beta-globin gene

Eleven variants (SNPs) were found in the study population (Table 1). These include rs713040, rs33930165, rs334, rs111851677, rs10768683, rs7480526, rs7946748, rs1609812, rs537944366, rs113152027, and rs33915217). Some of the SNPs in the beta globin gene are in linkage disequilibrium. Worthy of note are the relationships between rs1609812, rs7946748, rs10768683and rs713040, with D’ of 0·824 – 1·0 and r^2^ of 0·508 – 0·887 for each pair of the SNPs (Table 2). Only three of the SNPs (rs713040, rs10768683 and rs1609812) were found in all the SCD patients. The rs33915217 SNP was found in one patient in the separate SCD cohort.

**Table 1:** SNPs seen within the beta-globin gene, their frequencies and significance.

| SNP rs# | (HGVS nomenclature) | Allele frequency in cases (2n = 246) | Allele frequency in controls (2n = 234) | Allele frequency in YRI (1000 Genomes Phase 3 Project) | Allele frequency in Africans (1000 Genomes Phase 3 Project) | Global Allele frequency (1000 Genomes Phase 3 Project) | ClinVar annotation |
| --- | --- | --- | --- | --- | --- | --- | --- |
| rs713040 | NC_000011.10:g.5227013A>G | 1.000 | 0.892 | 0.894 | 0.884 | 0.714 | Synonymous mutation |
| rs33930165 | NC_000011.10:g.5227003C>T | 0.058 | 0.018 (n=4, heterozygous) | 0.028 | 0.013 | 0.003 | Missense variant HbC, pathogenic |
| rs334 | NC_000011.10:g.5227002T>A | 0.942 | 0.085 (n=20, heterozygous) | 0.139 | 0.100 | 0.027 | Missense variant HbS, pathogenic |
| rs33915217 | NC_000011.10:g.5226925C>A |  | - | 0.000 | 0.000 | 0.000 | Beta thal+, beta thal major |
| rs111851677 | NC_000011.10:g.5226822A>G | - | 0.014 (n=3, heterozygous) | 0.019 | 0.022 | 0.006 | Beta thal benign* (conflicting submission) |
| rs10768683 | NC_000011.10:g.5226561C>G | 1.000 | 0.892 | 0.894 | 0.890 | 0.720 | Intron variant benign |
| rs7480526 | NC_000011.10:g.5226503A>C | - | 0.256 | 0.338 | 0.379 | 0.369 | Intron variant benign |
| rs7946748 | NC_000011.10:g.5226496G>A | - | 0.119 | 0.065 | 0.065 | 0.099 | Intron variant benign |
| rs113152027 | NC_000011.10:g.5226357G>A | - | 0.026 | 0.009 | 0.007 | 0.002 | Intron variant benign |
| rs1609812 | NC_000011.10:g.5225911G>A | 1.000 | 0.889 | 0.894 | 0.884 | 0.714 | Intron variant benign beta thal |
| rs537944366 | NC_000011.10:g.5225542T>C | - | 0.0043 (n=1, heterozygous) | - | 0.002 | 0.0004 | 3' UTR variation uncertain significance |

YRI – Yoruba in Ibadan (1000 Genomes Phase 3 project)

**Table 2:** D’ and r^2^ values between the observed SNPs in the beta-globin gene as generated by Haploview 4·2.

| D' values |  |  |  |  |  |  |  |  |  |
| --- | --- | --- | --- | --- | --- | --- | --- | --- | --- |
|  | rs537944366 | rs1609812 | rs113152027 | rs7946748 | rs7480526 | rs10768683 | rs111851677 | rs334 | rs33930165 |
| rs1609812 | 1.0 |  |  |  |  |  |  |  |  |
| rs113152027 | 1.0 | 1.0 |  |  |  |  |  |  |  |
| rs7946748 | 1.0 | 0.824 | 1.0 |  |  |  |  |  |  |
| rs7480526 | 1.0 | 1.0 | 0.48 | 1.0 |  |  |  |  |  |
| rs10768683 | 1.0 | 0.955 | 1.0 | 1.0 | 1.0 |  |  |  |  |
| rs111851677 | 1.0 | 0.043 | 0.478 | 0.102 | 1.0 | 0.0 |  |  |  |
| rs334 | 0.05 | 0.62 | 0.28 | 0.36 | 1.0 | 0.06 | 1.0 |  |  |
| rs33930165 | 1.0 | 0.18 | 1.0 | 1.0 | 1.0 | 1.0 | 1.0 | 1.0 |  |
| rs713040 | 1.0 | 1.0 | 1.0 | 1.0 | 1.0 | 0.96 | 0.011 | 0.379 | 0.412 |
| r<sup>2</sup> values |  |  |  |  |  |  |  |  |  |
|  | rs537944366 | rs1609812 | rs113152027 | rs7946748 | rs7480526 | rs10768683 | rs111851677 | rs334 | rs33930165 |
| rs1609812 | 0.001 |  |  |  |  |  |  |  |  |
| rs113152027 | 0.0 | 0.003 |  |  |  |  |  |  |  |
| rs7946748 | 0.0 | 0.508 | 0.002 |  |  |  |  |  |  |
| rs7480526 | 0.008 | 0.064 | 0.011 | 0.048 |  |  |  |  |  |
| rs10768683 | 0.001 | 0.776 | 0.004 | 0.636 | 0.076 |  |  |  |  |
| rs111851677 | 0.0 | 0.0 | 0.151 | 0.002 | 0.032 | 0.0 |  |  |  |
| rs334 | 0.04 | 0.04 | 0.05 | 0.07 | 0.37 | 0.0 | 0.05 |  |  |
| rs33930165 | 0.0 | 0.0 | 0.001 | 0.002 | 0.011 | 0.003 | 0.0 | 0.05 |  |
| rs713040 | 0.0 | 1.0 | 0.004 | 0.676 | 0.073 | 0.887 | 0.0 | 0.002 | 0.001 |

### Prevalence of beta thalassemia in the study population

No beta thalassemia mutation was found in the SCD patients of this study main cohort, while a mutation that has been associated with benign beta thalassemia (rs111851677) was found in three individuals in the control group in the heterozygous state at a frequency of 0.014. These three individuals had normal haematocrit >36%, HbF levels <1%, and HbA_2_ levels <3%. One of the individuals had a low MCV of 76.8% and an MCH of 23.8. One of the SCD patients in the separate cohort was observed to have a moderately severe beta thalassemia intermedia mutation (rs33915217C>A) (Supplementary Figure 3). This SNP is an intronic variant at chr11:5,226,925 (hg38). The hematological parameters of this patient with beta thalassemia mutation are also shown in Supplementary Figure 3.

### SNP haplotypes and their relationship

Eight haplotypes were found in this study population (Figure 1). Two distinct ancestral haplotypes were discovered in this study population which comprised of about 88% of Yoruba indigenes from Oyo, Lagos, Ekiti, Ogun, Ondo, Osun, and Kwara States. The other individuals had one of Edo, Imo, Anambra, Abia, Enugu, Delta, Kogi and Benue as their states of origin (paternal). The major ancestral haplotype (TAGGAGATCG) occurred at a total frequency of about 0·850 and the minor ancestral haplotype (TGGAACATCA) occurred at a total frequency of 0·116 (Table 3). Four SNPs (rs713040, rs10768683, rs7946748 and rs1609812) differentiate the major and minor ancestral haplotypes, and they are in linkage disequilibrium (Table 2). Only one haplotype (TAGGAGAACG), a variant of the major ancestral haplotype, was found in all the participants with SCD in the homozygous state (HbSS) and another variant (TAGGAGATTG) was associated with haemoglobin C (HbC) (found in those with HbSC and HbAC genotype). This SCD haplotype was not peculiar to the Yoruba people alone, as eleven (8.4%) of these participants with SCD, had both paternal and maternal states of origin outside the Yoruba states. The minor ancestral haplotype and its variants were found in twenty-seven (24%) control participants in the heterozygous state, no participant had the homozygous state

**Figure 1:**
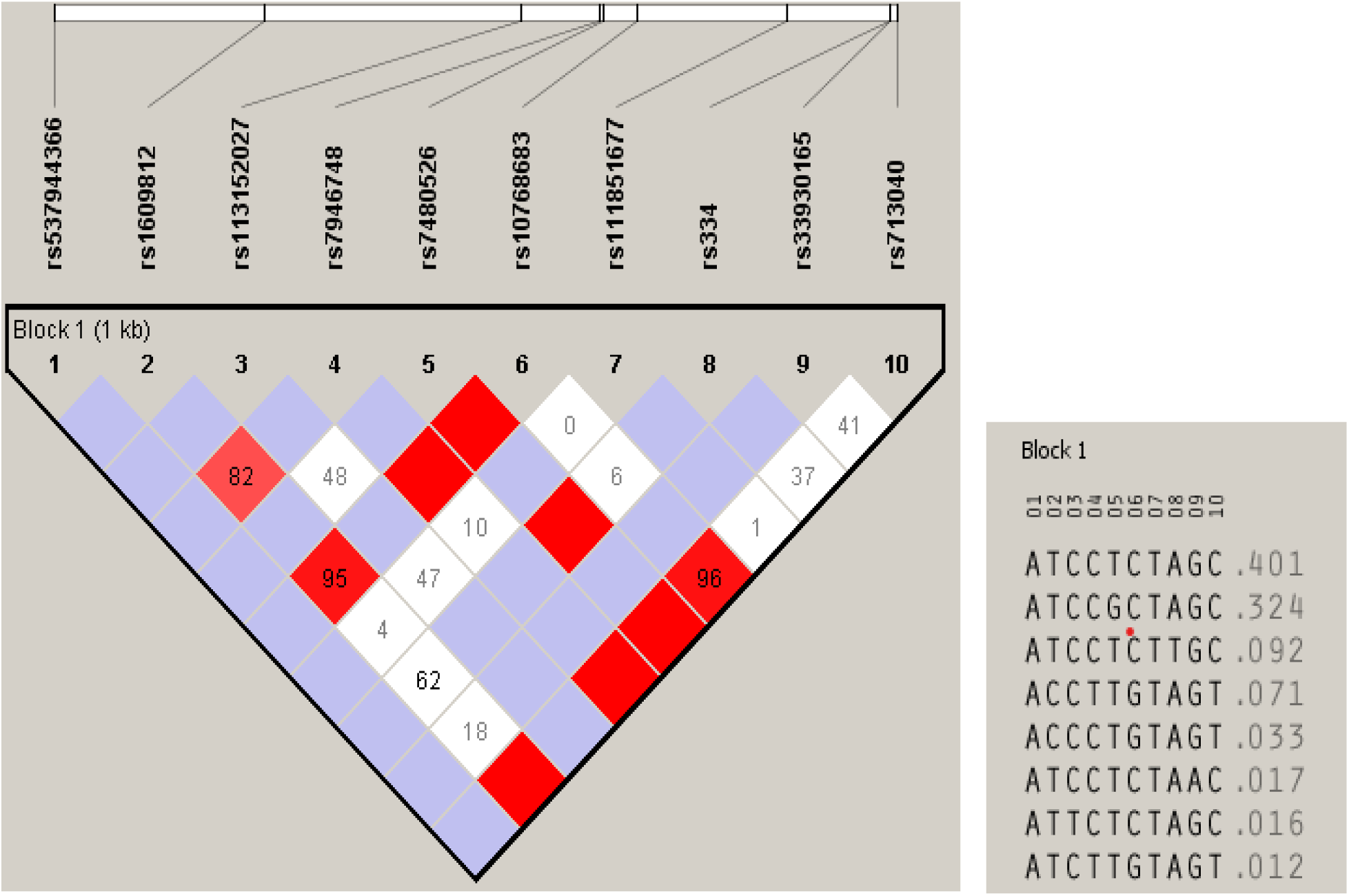
Haplotypes found within the beta globin gene, their frequencies and relationships generated by Haploview 4.2. Boxes shaded red show the strongest relationship between SNP alleles. Numbers in boxes = D’ values between two linked positions White boxes = LOD <2 and D’ <1 Purple boxes = LOD <2 and D’=1 Red boxes = LOD ≥ 2 and D’ = 1 (LOD = logarithm of the odds, D’ = relative linkage disequilibrium).

**Table 3:**
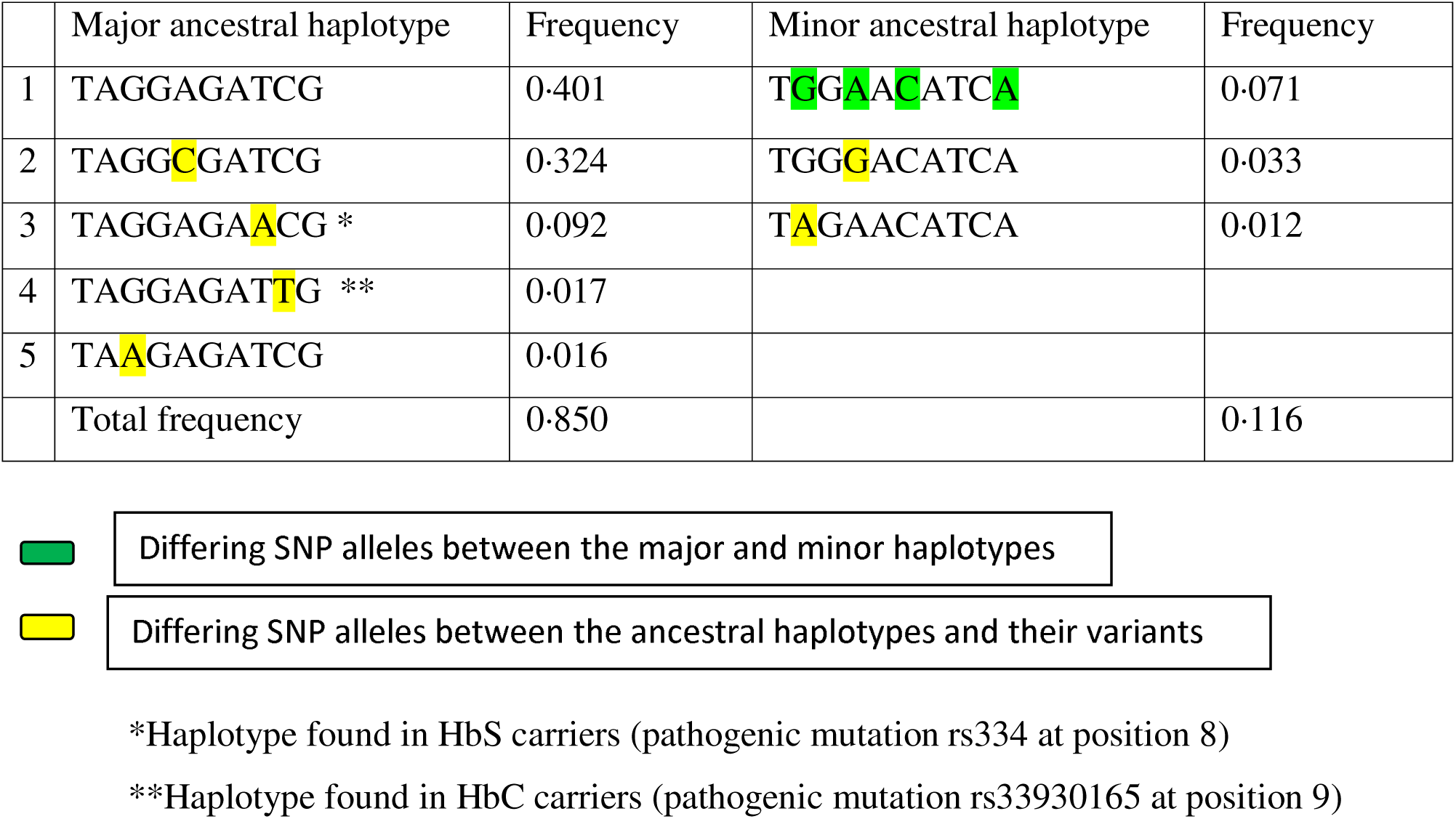
Partitioning of haplotypes into major and minor ancestral haplotypes and SNP allele differences between the haplotypes.

## DISCUSSION

The beta-globin gene accommodates a lot of mutations, some of them leading to hemoglobinopathies. The hemoglobinopathies are regionally specific, as a unique combination of genetic mutations in the globin genes are found in each population^8,23^. Beta thalassemia mutations are due to about 300 mutations in the globin gene^24^. None of these beta thalassemia mutations was found in our main study population of 240 individuals comprising of 123 SCD patients and 117 individuals without SCD. The only individual we have found with a moderately severe beta thalassemia intermedia mutation was an SCD patient in the separate cohort. This mutation is rs33915217C>A. There are three alleles associated with this SNP. The other two are rs33915217C>G and rs33915217C>T^3^. The SNP is a rare variant but the last two alleles are more common than the one found in this study^19^. The three mutation alleles cause severe form of beta thalassemia media by reducing the efficiency of splicing of the normal 5’ splicing site, leading to reduction of full-length mRNA^3,25^. Two patients, one from Northern Europe and the other from the Mediterranean region have been found with the A allele in the heterozygous and homozygous states respectively^25^. The hematological parameters of the patient with beta thalassemia in this study (low HbA, high HbF and HbA_2_ with low MCV), are also comparable with patients in which the G allele has been found^3^. To the best of our knowledge, this is the first report of this variant (rs33915217C>A) in the African population and in an SCD patient. This very rare allele was not found in the 1000 Genomes Phase 3 project, but has been reported in a non-Finnish European in the gnomAD exome v4 dataset (gnomADe:nfe) at a very low frequency of 0.000009^21^.

This result confirms previous reports^11,15^ from studied samples in Western and Northern Nigeria respectively; that beta thalassemia is prevalent at a very low frequency of <1% in Nigeria. This is in contrast to previous reports^9,10^ that beta thalassemia is at a higher prevalence of about 20 – 25% in non-SCD individuals and about 6 – 10% within SCD patients^14,26^. The discrepancies in these reports and findings in this study is because those studies relied solely on the measurement by HPLC, of HbA_2_ greater than 3.5% and sometimes, HbF >1% inclusive to determine the prevalence of beta thalassemia in the Nigerian population. Esan^11^ also measured HbA_2_ levels to determine the prevalence of beta thalassemia in Western Nigerians. He adopted the starch-block electrophoresis method and had only few people with HbA_2_ levels above 3.8%; and hence his conclusion of low prevalence of beta thalassemia in the population. Using HPLC, some of the patients in our study cohort also had elevated levels of HbA_2_ above 3.5%, but genetic analysis showed no beta thalassemia mutation. Inusa et al^15^ also reported cases of individuals with elevated HbA_2_, but with no beta thalassemia on genetic confirmation. HbA_2_ levels could also be elevated in individuals with alpha thalassemia^16^.

For our study population, asides the HbS (rs334) and HbC (rs33930165) mutations, the remaining SNPs observed are intronic variants. The rs713040G is an alternate allele and has a higher global allele frequency than the reference allele, rs713040A. The frequency in the study control population, the African population and global population is almost the same at > 0.800. However, this alternate allele was found in individuals with SCD in this study at a frequency of 1.000. The rs33930165T codes for the C haemoglobin and so far, it has only been found to be prevalent in some African populations at a total frequency of 0.013 and in the YRI at 0.028^21^. The frequency of the HbC mutation in the control group of this study is 0.018. The rs334A occurs in the same codon, but the next nucleotide base to rs33930165. It also has the highest frequency in African populations at a frequency of 0.100 which is not too different from what was observed in our study control population (0.085) but lower than that found in YRI (0.139) (p<0.001). This variant when inherited in the homozygous state causes SCD. This variant may also be inherited in the heterozygous form with the rs33930165 variant (Hemoglobin C) to give a milder form of SCD, known as HbSC^27,28^.

The rs111851677 is an intron variant in the intervening sequence I (IVS-I-108) of the beta-globin gene. Its frequency in the study population is low at 0.014. The global allele frequency (GAF) is 0.006 and it is most prevalent in the African population at a frequency of 0·022 and in the YRI at 0.019. It has its highest prevalence in the ASW (African Ancestry in Southwest US, 0·049)^21^. This allele was recorded as a novel mutation in a beta thalassemia carrier in Cuba^29^ and in an Iranian^30^, and later reported in the Greek population in 2008^31^. There have been conflicting submissions about its clinical significance in the population. It has been suggested to be associated with benign beta thalassemia, especially when inherited in compound heterozygosity with another defect on the beta globin gene, but its precise role is still unclear^30,32^. It has been suggested^29^, that due to the nucleotide’s proximity to the 3’ splice junction, and close to the branch site, the mutation may lead to ineffective RNA splicing, leading to beta thalassemia. However, when the red blood cell parameters of the three individuals with this mutation in the study sample were checked, there were no signs of beta thalassemia. There was normal haematocrit >36%, HbF levels were <1%, and HbA_2_ levels were also <3%. One of the individuals had a low MCV of 76.8% and an MCH of 23.8 which are most likely due to alpha thalassemia, which has a high prevalence of about 35% - 45% in the population^16,18^. The very high prevalence of alpha thalassemia might not have supported the penetration of beta thalassemia mutations, and hence, its very low frequency within this study population.

The rs10768683G is a benign intron variant with frequency in our study population (0.892) the same as those in YRI (0.894) and Africa (0.890) and global allele frequency is lower at 0.720. The rs7480526C is a benign intron variant but its frequency in the study participants (0.256) is lower than that found in YRI (0.338), in Africans (0.379) and across the globe (0.369) (p=0.003). rs7946748A is also a benign intron variant with frequency in the study control population (0.119) close to that of the global allele frequency (0.099) and higher than that found in the YRI and African populations (0.065) (p<0·001). rs113152027, a benign intron variant was found at a frequency of 0.026 in our study population, a frequency much higher than that found in the general African population (0·0050) and across the globe (0·0016) (p<0·001). This variant is most prevalent in individuals of African Ancestry and PUR (Pueto Ricans in Pueto Rico) at a very low frequency of < 0·01 except in the LWK (Luhya in Webuya, Kenya) where a frequency of 0·02 has been reported^21^.

rs1609812 (C>T) is another benign intron variant with the frequency in the study population (0.889) close to that of the frequency in YRI (0.894), in African populations (0.884) and higher than the global frequency (0.714) (p<0·001). rs537944366 (T>C) is a very rare variant found in this study in only one individual in the heterozygous state (frequency of 0.0043). It is a rare 3’ UTR variant with uncertain significance. The global (0.0003) and African frequency (0·002) are also very low as found in the study population. This variant was not seen in YRI of the 1000 Genomes Phase 3 project and has only been reported in MSL (Mende in Sierra Leone) (0·012) and in gnomADg:nfe (Non-Finnish European population) (0.00003) in the whole world^21^. This variant is being reported for the first time in the Yoruba people of Nigeria. An analysis of the variants seen in the HBB gene (exons 1 and 2) of a Senegalese population showed that all the variants seen in this study population were also observed. In addition to these, SNPs such as rs35209591 (frequency >0·03), rs33945777, rs34598529, rs72561473 and rs33944208 (frequency <0·03) were also found^33^.

Two ancestral haplotypes; a major haplotype with a frequency of 0.850 and a minor haplotype with a frequency of 0.116, were discovered in this study population. Four SNPs in linkage disequilibrium differentiate these two ancestral haplotypes: rs713040, rs10768683, rs7946748 and rs1609812. The alleles of SNPs (rs713040G, rs10768683G and rs1609812A) found on the major haplotype are the most frequently occurring allele globally (about 0·72) and have their highest frequencies in Africa (0·89), then Europe (0·83), South Asia (0·58) and least frequent in East Asians (0·51). This finding supports the origin of man in Africa and his migration out of the continent towards Europe and Asia.

The other alleles of these SNPs (rs713040A, rs10768683C and rs1609812G) are most prevalent in the EAS (East Asian) and SAS (South Asian) populations. They occur in EAS and SAS at a frequency of about 0·49 and 0·42 respectively. rs7946748A has its highest frequency in SAS (0·197) and the least frequency in EAS (0·025), with a global frequency of 0·099^21^. This suggests that this mutation likely arose in the SAS population. The highest prevalence in the South Asian regions, of these four SNP alleles on which the minor ancestral haplotype differs from the major ancestral haplotype alludes to the fact that individuals from around this region might have migrated and mixed with the indigenous African population some time in the past.

We conclude that the major ancestral haplotype is typical of Africa, while the minor ancestral haplotype is atypical of Africa. The prevalence of the minor ancestral haplotype in this population alludes to genetic admixture from an immigrant population. The homozygous state for the minor ancestral haplotype was not observed in this study population. According to Hardy Weinberg equilibrium, the homozygous state for the minor ancestral haplotypes with a population frequency of 0·116 is expected at 0·014; which this study sample size may not have the power to detect. The major ancestral haplotype and one of its variants have been found in a Turkish population (as haplotype II & I respectively) at a frequency of 0.727 and the minor ancestral haplotype and its variants (as haplotype III - VII) at a frequency of 0.273^34^.

Only one haplotype, the major ancestral haplotype (TAGGAGAACG) was associated with the sickle and haemoglobin C mutations in this population. The SCD patient in whom the beta thalassemia intermedia mutation was found in this study is a heterozygote for the two ancestral haplotypes. Hence the beta thalassemia mutation is associated with the minor ancestral haplotype. The sickle and HbC haplotypes are variants of the major ancestral haplotype (TAGGAGATCG). The sickle and HbC mutations in this population arose on the major ancestral haplotype (rs334 and rs33930165 at position 8 and 9 respectively) (Table 3) and have undergone no or low mutation since then. This points to the fact that there are very low recombination events within the beta-globin chain and single nucleotide polymorphisms seen in the beta-globin gene of this population are most likely due to spontaneous mutations. However, in the Turkish population mentioned above^34^, 60% of the sickle mutation was found on haplotype II, which is the major ancestral haplotype found in this study and 10% was found on haplotype III, the minor ancestral haplotype found in this study. Haplotype I, the major haplotype found in the Turkish population (frequency, 0·481), and a variant of the major ancestral haplotype found in this study (TAGGCGATCG, frequency 0·324), was associated with 20% of the SCD cases. This shows that the sickle mutation in most of the Turkish population arose on the major ancestral haplotype found in this study, but unlike in this study population, the sickle haplotype has undergone some mutation in the Turkish population. The HbS mutation found on the minor ancestral haplotype in the Turkish population confirms the independent origin of the HbS mutation outside of Africa^35^, most probably that of the Arab-Indian origin.

For the Senegalese study^33^, six SNPs found in the population were not observed in this study. Five of the SNPs were at a frequency lower than 0·03, while only one (rs35209591) was found at a higher frequency. The larger sample size used in the Senegalese study may have enhanced the detection of the five minor allele SNPs not found in this study. On the other hand, the differing set of SNPs may confirm a different beta-globin gene haplotype in the Senegalese population (the Senegal haplotype). Seven of these SNPs in the YRI population of the 1000 Genomes Phase 3 project were analysed for linkage disequilibrium^36^ with similar results to what we were able to generate with Haploview; affirming the accuracy of our sequencing results, analysis and deductions.

## CONCLUSION

Beta thalassemia is at a low prevalence in this study population and it is associated with the minor ancestral haplotype. There is a need to re-assess the diagnosis of beta thalassemia in this and other African populations.

## Data Availability

All data produced in the present work are contained in the manuscript and supplementary materials. Further information needed are available upon reasonable request to the authors

## ACKNOWLEDGEMENTS

Doris Duke Charitable Foundation

Center for Global Health, University of Chicago

