## Supplemental Table 1, Supplemental Figure 1,2,3 for "The genetic architecture of the β-globin chain in individuals with and without sickle cell disease in Nigeria: A case for beta thalassemia?"

**SUPPLEMENTARY MATERIALS**

Table 1: Primers used for human beta-globin gene amplification

| # | Primer Name | Primer sequence (5’ – 3’) | Length of amplicon |
| --- | --- | --- | --- |
| 1 | Forward Primer 1 (F1) | AACTCCTAAGCCAGTGCCAGAAGA |  |
| 2 | Reverse Primer 1 (R1) | TCATTCGTCTGTTTCCCATTCTAAAC | 774 bp ^37^ |
| 3 | Forward Primer 2 (F2) | ATGTTTTCTTTCCCCTTCTTTTC |  |
| 4 | Reverse Primer 2 (R2) | CAGAGATATTGCTATTGCCTTAACC | 649 bp (primer set 2 designed using Primer3) |
| 5 | Forward Primer 3 (F3) | CATGCCTCTTTGCACCATTCT |  |
| 6 | Reverse Primer 3 (R3) | CACTGACCTCCCACATTCCCTTTT | 574 bp ^37^ |

F: forward, R: reverse

| 5’ UT | EX 1 | IVS 1 | EX 2 | IVS 2 | EX 3 | 3’ UT |
| --- | --- | --- | --- | --- | --- | --- |

AAAAA

----------------------774 bp------------------------------------- -----------574 bp------------------------------

--------------------649 bp--------------------

Figure 1: Regions of the beta-globin gene amplified by each set of primers (Modified from ^37^

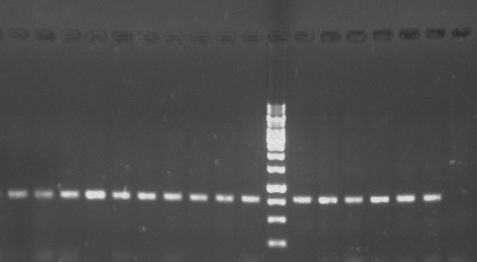

1000 bp marker

774bp

250 bp marker

Amplicon 1

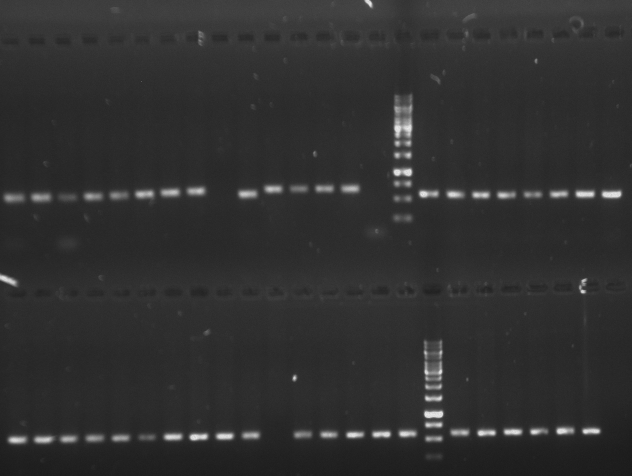

250 bp marker

574 bp

250 bp marker

649 bp

Amplicon 3

Amplicon 2

1000 bp marker

1000 bp marker

Figure 2: Gels showing the beta-globin gene amplicons from the three different set of primers.

Upper gel shows amplicons from F1R1(774 bp) and the lower gel shows amplicons from F2R2 (649 bp) and F3R3 (574 bp).

rs33915217

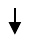

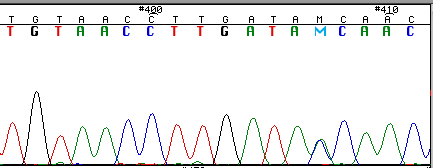

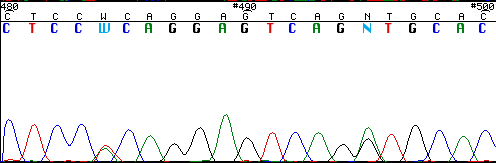

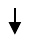

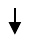

rs713040

rs334

Haematological parameters of patient with moderately severe beta thalassemia intermedia:

HbA	- 8%	HGB	- 9·5 g/dL	RBC	- 4·42

HbA_2_	- 4·8%	HCT	- 30%	RDW	- 22·4

HbF	- 11·9%	MCV	- 67·9	WBC	- 8·5

HbS	- 71·4%	MCH	- 21·5	PLT	- 349·5

Figure 3: Chromatogram of a patient with sickle beta thalassemia (Sβ^+^). rs334 is the sickle mutation and rs33915217 is the moderately severe beta thalassemia intermedia mutation.
